# Central desks as an approach for health governance in conflict settings – case studies from northwest Syria

**DOI:** 10.1101/2022.09.23.22280280

**Authors:** Maher Al Aref, Zedoun Al Zoubi, Munzer Al Khalil, Orwa Al Abdulla, Abdulkarim Ekzayez

**Author notes:** Corresponding author Corresponding author: Abdulkarim Ekzayez, MD MSc, Department of War Studies, King’s College London, Room K6.08, King’s Building, Strand Campus, Strand, WC2R 2LS, E, – Ext: 89913.

## Abstract

**Background:** The conflict in Syria affected severely the health sector; health infrastructure was damaged, the Damascus ministry of health withdrew from opposition held areas, health workers fled the country, and there has been always a shortage of funding and medical supplies. To address these needs, Syrian NGOs, INGOs, donors, and UN Agencies have been providing health interventions through humanitarian channels. However, many of these interventions were short termed, and there was no governance framework to guide the newly introduced parallel system, leaving it subject to individual organizations’ strategies and approaches.

To counter these challenges, local communities and Syrian NGOs established new platforms to govern and coordinate certain aspects of the health sector. These platforms are called “central desks”, which are perceived to be independent and neutral structures and can coordinate services between all actors. Examples of these structures are Syrian Immunization Group (SIG), Health Information System (HIS), the Infection Protection and Control initiative (IPC), and the Referral System network.

**Methods:** The research was based on an institutional approach to governance as presented by (Abimbola et. Al, 2017) and (Baez-Carmago and Jacobs, 2011) of health governance. We have investigated the central desks across the main themes; governance inputs of these central desks, such as strategic vision and legitimacy; governance processes, such as accountability and transparency, and governance outcomes, such as effectiveness and efficiency. Further to intensive literature review, eight focus group discussion were conducted, average pf 12 participants. Key themes then were deducted and coded. The qualitative analysis was done using NVIVO 12 software.

**Conclusion:** Central desks, that are not part of national ministries of health, are new innovative approaches that can increase the efficiency of health interventions in conflict settings. The detailed features of such desks should be context specific and locally informed and led.

## Introduction

Historically, global health can be rooted in the period of European colonialism, when health interventions were merely focused on securing colonizers from locally prevalent diseases. This is when western institutions studying “tropical diseases” flourished. However, gradually there were concerns that health threats can cross national borders, and if a harmful pathogen was left to spread in a low-income country, this pathogen can threaten the whole world. Throughout the years, therefore, the mission of global health expanded from focusing on tropical medicine to improving health equity worldwide and even strengthening local systems to fight diseases before it spread [1].

However, some could argue that the unequal balance of power in times of colonialism, when global health emerged, was having, and still having, great impact on unequal relations and inequalities in global health until today. Additionally, the various local political systems sometimes interfere with these efforts of global health in a way that can paralyse these efforts [2]. These political interferences can be very profound in times of conflict and political instability. Local governments, non-state armed groups, and other warrying parties usually interfere with provision of health services, which result in severe disruption of health governance in such settings. This pushed various actors to think of other ways and approaches to deliver health services and implement local health system strengthening interventions without being paralysed with such interferences.

Although several frameworks for health governance were developed [3], all of these frameworks were developed for stable settings with very limited applicability in armed conflict settings. For example the institutional analysis framework developed by Abimbola 2017 assumes the existence of functioning institutions to explore health governance [4]. Similarly, the World Health Organization (WHO) frameworks for health governance do have this underlying assumption [5]. When approaching health governance in conflict settings, therefore, it is important to use combinations of these frameworks and think of innovative solutions for how to navigate through conflict sensitivities and geopolitics.

There have been some examples of central projects approach to navigate through complicated political governance in fragile settings to implement health interventions. Key features of such central projects include independence, autonomous structure, and technical focus. In 2001, the fight of the “Big Three”, which are HIV, Tuberculosis, and Malaria, was a practical test on how to navigate through political and economic complexity in fragile and conflict affected countries in Africa. The fight required wide scale local, regional, and international collaborations and partnerships between donors, health organisations, ministries of health, and local communities. Two decades after the launch of this program, these three diseases have been eliminated in many of the targeted communities. It was not possible to achieve this without re-thinking global health structures and governance mechanisms [6]. Richard Feechem, who was the Managing Director of the Global Fund for Fighting AIDS, Tuberculosis and Malaria, believes that the Global Fund’s autonomous, multi-sectoral, technically-focused design was a key factor behind avoiding political complications and achieving better health outcomes [7]. Basically, the project established a parallel governance specifically designed to address this very health issue of these three diseases. This governance was cross national borders, very technical focused, and quite standardises across localities. The local engagement of this structure was done on communities level rather than political level allowing the project to involve local communities while avoiding domestic politics. Indeed, one of the conditions for a country to receive grants under this project was to allow this program to have an autonomous structure with high levels of independence.

However, the Global Fund project on HIV, TB and Malaria can be prescribed as a vertical project with minimum investment in and strengthening of local health system. In most countries, where these projects are implemented, these three diseases receive many more resources than other health threats or other components of the local health systems. It can be argued that these investments can have much larger returns in relation to improving health outcomes if channelled in a way that further strengthen local systems [8]. Furthermore, these central projects assume a level of functionality in national health systems, which might not exist in conflict settings.

To overcome this challenge, some organisations established new central structures that act as governing bodies for specific projects. The structure for the world bank project on HIV in Africa engages with a central national council that act as a governing body for the whole project and oversee the implementing partners – that might include ministries of health, NGOs, and grassroot organisations [9]. Such structures with a sense of centrality, that is merely focus on the technical nature of such projects, can help actors avoid political interferences while ensuring central coordination and leadership.**(Figure 1)**.

**Figure 1.**
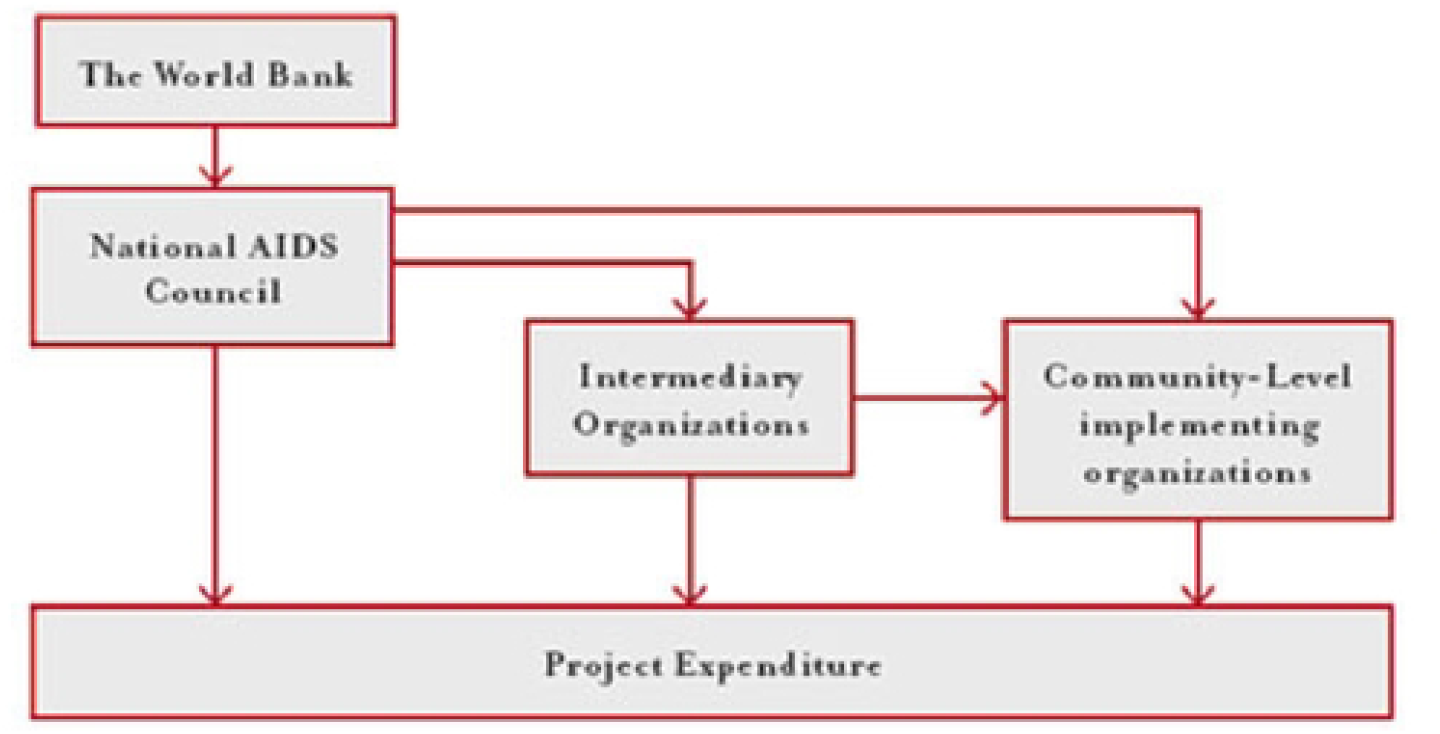
World bank - HN Program structure (World Bank HIV/AIDS Program, 2005)

Such projects with central technical structures, nevertheless, have some gaps and shortages in relation to the design and the functionality of these central structures that might result in poor implementation and poor health outcomes [10]. reviewed the effectiveness of the central HIV project in developing countries and they found various gaps that can be regarded as key systems constraints to scaling up of HIV treatment. These gaps include poor distribution of resources, low remuneration and accelerated migration of skilled health workers, verticalization of HIV treatment and its impact on disruption of national stewardship. They propose few recommendations to overcome these shortcomings. These recommendations include the need to evaluate the roles and the structures of community health workers in relation to HIV treatment access, international actions on brain drain, and greater investment in national human resource functions of planning, production, remuneration and management.

In conflict settings much attention is usually paid to life saving health interventions at the expense of health governance which receive little attention. The assumption in such settings is that conflict disrupt local health systems with different levels of collapse in health leadership. This might push humanitarian actors to establish a parallel system to coordinate health response. Such parallel system sometimes disrupts locally developed systems, which makes coordination and collaborations even more challenging. One way to navigate through these parallel systems could be through establishing central technical structures like the examples mentioned above. However, there is a high likelihood for political and military interferences that might make these central structures paralysed. There is a need, therefore, to design these technical structures carefully to avoid theses interferences while ensuring buy-in from all relevant actors.

The approach of central technical structures to navigate through complex political and sometimes militarised health governance is quite recent and still requires further development. In the Syrian conflict there were few examples of innovative approaches to establish and maintain such technical central structures in the context of northwest Syria. Our paper investigates the experience of four prominent central projects in northwest Syria, which are: the Health Information System Unit (HIS unit), the Syrian Board for Medical Specialties (SBOMS), the Referral central desk, and the central desk for Infection Protection and Control (IPC). Learning from these cases would help develop this approach of central technical structures further to be used as a tool to navigate through complex health governance in conflict settings.

## Methodology

The concept of central desks in the health sector in humanitarian settings is quite unexplored. Our study started with a targeted literature review for academic and grey literature to identify relevant resources and/or practical examples of this concept. The literature review included four major medical databases (Medline, Embase, Global Health, and Web of Science). The search strategy included three components; one for “central desks” with 8 search terms such as “central projects”; another for “humanitarian settings” with 10 search terms; and a third component for “health” with 14 search terms. The inclusion criteria were very wide to include any study with a focus on health projects with elements of centrality either on management levels or coordination levels. However, this search strategy returned only 4 studies. We used then targeted web search to look into grey literature from UN agencies websites, humanitarian websites, and NGOs reports.

The literature review was then followed with 8 focus group discussions conducted in Mersin/Turkey in November 2021. The participants included health policy makers from governmental and non-governmental health organisations operating in northwest Syria. Each FGD had an average of 12 participants and took about 2 hours. The FGDs were conducted and transcribed in Arabic. The transcripts were later translated and analysed in English The key themes then were deducted and coded. The qualitative analysis was done using NVIVO 12 software.

For the purpose of this paper, we define central desks as “technical health bodies that are established to govern and coordinate certain aspects of the health sector. These bodies are independent and neutral structures and can coordinate services between all actors”. We used a combination of two governance frameworks as developed by (Abimbola et. Al, 2017) and (Baez-Carmago and Jacobs, 2011) [4,13] to explore such central desks in the Syrian context.

Lastly, the study authors are all health practitioners and academics with extensive involvement in the health response in Syria. We thus used our accumulative experience and observations to identify relevant experiences and interpret some of the findings. This interpretation was also validated through a network of health experts in the context of northwest Syria.

### The evolvement of central desks in northwest Syria

Since the withdrawal of the Damascus Ministry of Health (MoH) from opposition-held areas in northwest Syria, the health governance in these areas went through four major stages. The first could be characterized as grassroots, fully decentralized, loosely connected governance. During this stage activists and health workers coordinated their work with horizontal networks and local bodies to respond to the medical needs resulting from the Syrian government violence. There have been, however, examples of innovative bottom up approaches to establish locally led health bodies, such as the establishment of Idleb Health Directorate (IHD) in 2012 [14,15].

The second stage started with forming the opposition Syrian Interim Government (SIG) in March 2013. The SIG tried to form structures similar to the Damascus government. A total of eight Health Directorates (HDs) were formed. This stage ended early 2015, when the relationships between HDs and SIG deteriorated due to poor planning and coordination, competing mandates, inefficiencies in allocating and using funds, as well as to the fact that SIG was formed in Turkey, whereas HDs were all inside Syria. This stage witnessed the formation of the first central desk at the end of 2014, which is the Syria Immunization Group. This was in response to the urgent vaccination needs with the emergence of Polio in late 2013, and the inability of the SIG to manage such national vaccination campaign. There was a need to establish an independent body that can govern and coordinate the vaccination activities across all actors including international and local NGOs. Initially, the executive arms of the Syrian Opposition Coalition SOC represented by the Assistance Coordination Unit (ACU) was responsible of leading this national campaign. However, due to political sensitives and the need to work with all actors, some of which cannot work directly with the opposition entities, the Syrian Immunization Group was established. In December 2015, SIG, ACU, HDs, and a group of Syrian NGOs issued a statement in which they considered the Syria Immunization Group as an independent and neutral body that is exclusively responsible for managing vaccination interventions in opposition held areas.

The third stage started in 2015 with forming alliances and coordination platforms among Syrian NGOs and the HDs. This encouraged donors, in particular the German government and the European Union, to allocate funds to HDs, and support capacity-building programs between 2017 and 2019. Aiming to encourage other health actors to join efforts, more central desks were established during this stage. This includes the central desks of drugs control, Health Information System (HIS), Syrian Board of Medical Specialities (SBOMS), the Referral System. All these central desks were independent from any political bodies with key roles of Syrian NGOs and HDs in maintaining these desks.

The fourth stage started in 2019 when the functionality of the HDs was drastically declined. This was after the Syrian regime took over large opposition held areas in the governorates of Daraa, Quneitra, Rural Damascus, Homs, Hama and Latakia; and after the expansion of Hayaat Tahrir Al-Sham (HTS), an Islamist militia, that became the dominant power in Idlib, putting its administrative arm, the Salvation Government, in charge of all services. In this stage, the humanitarian actors started to realise that the operational areas of HDs have shrunk, and the ability of the HDs to act as an exclusive health governance body in opposition held areas is under question in light of the dominance of HTS. This ability was also negatively affected by the cut of HDs funding by some international donors. In northern Aleppo, the operational capacity of Aleppo health directorate was also limited in light of the direct provision of services by the Turkish MoH in areas controlled by the Turkish government. Moreover, the presence of the HDs inside the country makes them subject to allegations about potential connections with the various militias and terrorist groups. Therefore, this stage was marked by a decline in the power of the remaining HDs, and some NGOs started to distance themselves from HDs.

Accordingly, humanitarian actors started to look for alternative structures that are resilient, agile, and independent such as the central desks. The level of autonomy of these central desks increased to overcome some of the obstacles associated with the health directorates. The relative success of the Syria Immunization Group, as the first central desk, made it an example to be followed. Consequently, the other central desks, such as SBOMS, HIS, and drugs control, started to gain more attention from NGOs, INGOs, and UN agencies – who started to build other central desks such as the referral system and the Infection Prevention and Control (IPC) central desk.

There are key features and practicalities that allowed these central desks to overcome the complex challenges of the health governance in northwest Syria. The leadership of these desks was usually based outside Syria, giving them relative immunity from accusations of any link with local militias or terrorist groups. Second, their independent structures gave them neutral positioning, allowing them to be supported by all NGOs and HDs. Third, their independence from each other, although caused inefficiencies sometimes, allowed them to mitigate risks; a failure in one central desk would not cause a failure from the others. Fourth, the political parties did not view these central desks as competing authorities, so they gave more operational space compared to the HDs. At the same time, NGOs did not view these bodies as affiliated with any political authorities, which allowed donors and humanitarian actors to engage largely with these bodies.

Today, considering the complex governance reality in the region, central desks are still an essential component in the health sector in northwest Syria, playing a critical coordinating and service provision roles working in coordination with health directorates, NGOs and the health cluster in Gaziantep.

### Case study 1: the Health Information Unit

The idea of central desks can be traced in the health system in Syria before the conflict. Some of the central medical services were not framed institutionally within the vertical system of the Damascus MoH [16–19]. One of the first central “project” to be initiated by Damascus MoH before the crisis was related to health information due to the MoH inability to provide necessary information to develop health plans [20].

Humanitarian responses in conflict settings requite data on mortality, morbidity, and health services to determine needs, design and evaluate interventions, and document conflict impact on health of population for operational and advocacy purposes [21]. This is usually done through rapid assessments and through existing data infrastructure. At a later stage in chronic crises, humanitarian health actors require more advanced data systems to collect, manage, and use health information. Health Information System (HIS) as defined by WHO is the “generation of information to enable decision-makers at all levels of the health system to identify problems and needs, make evidence-based decisions on health policy and allocate scarce resources optimally” [22]. Our paper will focus on Routine Health Information System (RHIS) which concerns health facilities data.

In Syria, the conflict severely affected the weak health information systems across the country [23]. With the start of the humanitarian response in northwest Syria NWS, the HIS was neither systematic nor standardized. In 2013-2014 Individual data collection systems were set up by various NGOs for their own programmatic needs, frequently collecting data that had not been defined or compiled using uniform standards and definitions. As a result, it was very complicated to monitor and evaluate the progress and achievement of humanitarian health programs towards the humanitarian objectives. Later, the District Health Information System DHIS was established and supported by NGOs and WHO for health programs in NWS [24].

The implementation of the DHIS software and the need to collect further facilities information necessitated a central coordination mechanism between the various actors involved in the provision of health services in NWS. Accordingly, the HIS unit was established in 2016 with a core team in Gaziantep, that works closely with the NGOs and the Health Cluster, and a field team that works closely with the HDs. Since 2016, the HIS unit has been playing a central role in coordinating the collection, management, and use of health information in the health sector in NWS. Main key achievements of the unite since its start-up were; the training of almost 250 health care providers and 200 data entries from around 165 health facilities in NWS, for basics of Health Information system and national health indicators according to the DIHS. The unit also contributed majorly at COVID 19 epidemic time by playing the lead role for supporting surveillance reports in partnership with the EWARN and the WHO covid 19 taskforce. Agreement on unified diseases coding system ICD 10, regular monthly report from PHC and Hospitals; also considered as a key achievement of HIS unit.

Based on the data analysis and findings from the FGDs, the current HIS unit for NWS lacks legitimacy due to several challenges. These challenges include: (1) weak cooperation from NGOs that run health facilities in the region as some actors underestimate the importance of the HIS unit; (2) poor internal regulations and operational procedures; and (3) limited technical support from WHO and the health cluster. The legitimacy of the HIS unit, according to the participants is gained from the following actors/factors i) NGOs, ii) the health cluster, iii) SIG MoH, iv) the quality of the HIS products, and v) WHO respectively.

One participant in the FGDs, Dr Mohamed Zahid; stated *“Quality of information should be the source of legitimacy, as long as the data collection and presentation methodology are accurate and unbiased the HIS unit would be a legitimate body in the health sector. In other words, there is a significant challenge to the HIS Unit to approach trustable and high-quality methods of data collection and analysis.”*.

The legitimacy of the HIS unit can be improved by several context-specific factors. The top two factors were quality of data products and the ability to cover wide geographical areas. 33% of the participants said that increasing the quality of the HIS reports and data products is the main source of legitimacy. 17% of them believed that the NGOs cooperation which is closely linked to extending the geographical coverage of the HIS unit contributes to promoting the unit’s legitimacy. Furthermore, it was believed that the outbreak of COVID-19 drew attention to the importance of reliable information during emergency and epidemics; and therefore, the demand for information increased which has raised the legitimacy of the HIS unit. **(Figure 2)**. One participant in the FGDs stated *“The COVID-19 outbreak promoted the legitimacy of the Unit. However, at the same time, the application of the HIS tools outside the COVID-19 context has been delayed.”*

**Figure 2:**
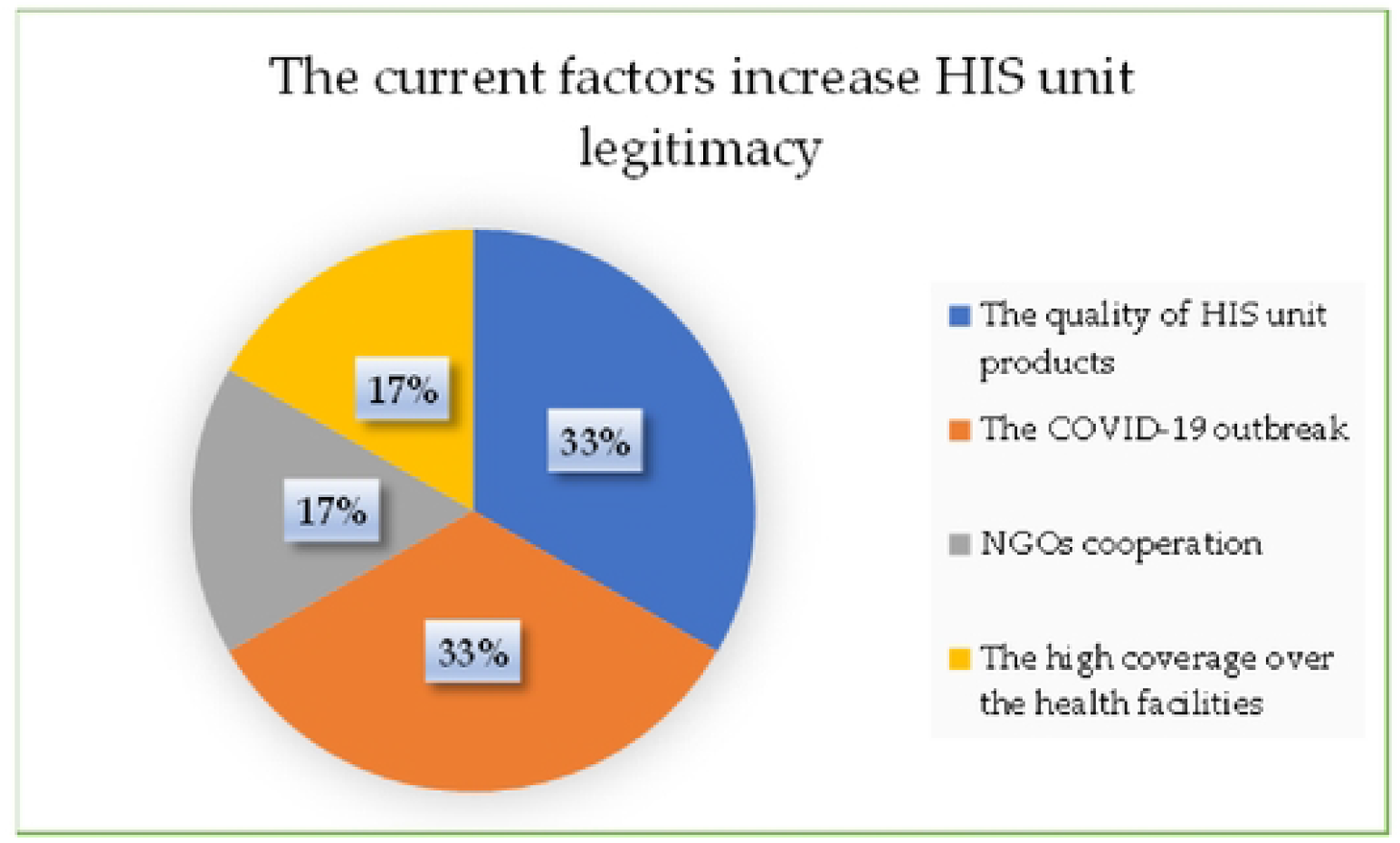
the current factors increase HIS unit legitimacy

The operational model of the HIS unit for NWS was investigated during the FGD to get better understanding of its roles and responsibilities besides how it contributes to the achievement of the humanitarian health objectives. According to the participants’ views, the current operational model of the HIS unit is considered to be rigorous to fulfil its technical and coordination roles. The main functions of the HIS unit include building the data infrastructure and reporting lines, producing facility level morbidity and mortality reports, and producing case level reports for COVID-19. the HIS unit has a vision which is to support all health actors with a holistic understanding of the principle public health indicators. However, the participants believed that this vision is not translated into clear strategy and operational procedures.

Accountability and transparency were reported to be key weaknesses of the HIS unit. The unit does not report to any governing body that is not involved in implementation, which is perceived by participants as lacking accountability enforcement. Additionally, there are no internal accountability mechanisms or procedures for transparency within the unit in relation to their ways of working, finance and resources, and implementation activities.

One participant, Dr Salah Safadi, stated *“There is no clear internal accountability process within the Unit neither upwards nor downwards. Of course, the change of government bodies played an important role in this ambiguity. Furthermore, accountability towards the beneficiaries, including health facilities, organizations, and healthcare providers, is still unclear.”*

Despite of the above-mentioned weaknesses and gaps, the participants agreed on the importance of having a central desk for health information. The agreed that the HIS unit should be further supported financially and technically to fulfil its roles and responsibilities in improving the availability and the quality of health information in the health sector in NWS.

### Case study 2: the Syrian Board of Medical Specialties (SBOMS)

Choosing the post-graduate specialty for Syria medical schools’ student, before 2011, was based on their scores in standardized medical exam for graduation besides their cumulative grades in the context of their desires [25]. Similar to many other middle-income countries (before the crisis), there was a lack in the post-graduate education and professional development in Syria [26]. It was not until 2012, the Syrian commission for medical specialties was established in Damascus through the legislative decree 68 to be the national body responsible of overseeing medical training programs [27].

The conflict in Syria pushed many health personnel to flee the country, which, coupled with the targeting of health workers, resulted in a remarkable gap in the number of skilled and specialized health staff [28]. There have been some Syrian-led initiatives in NWS to fill this gap. The most remarkable initiative is the SBOMS (Syrian Board of Medical Specialties), which was established in the middle of 2015 as a central desk that aims to supply the health labour market with new specialised doctors through resuming the training programs for medical graduates with standardised examination and certification consequently. SBOMS is loosely affiliated with the Syrian Interim Government’s Ministry of Health (SIG), and it collaborates with the health directorates to expand postgraduate and specialty opportunities for doctors in NWS based on projected healthcare system needs [29] [30]. SBOMS was established as an independent entity with legal and financial autonomous. SBOMS have 23 scientific committees (composed of specialists consultants inside Syria and expatriate Syrian doctors in the diaspora) that support post-graduate training, examination, and certification for a variety of specialties such as internal medicine, general surgery, vascular surgery, pediatric surgery, orthopedic surgery, urology, pediatrics, cardiothoracic, obstetrics and gynecology, ENT, ophthalmology, anesthesia and intensive care [29]. According to the information provided during the FGD; SBOMS managed to grant graduation for 204 specialized doctors tell November 2021 since the establishment, conducted 263 medical exams, and 7 specialized conferences along with launching multi-functional educational platform to provide medical training courses and lectures as the unit key achievements.

With regards to SBOMS legitimacy, 50% of the FGDs participants considered SBOMS as highly legitimate. They attribute this legitimacy to SBOMS critical role in filling gaps in the health labour market in the region, and to the advance quality of its education system which is evidence based and up to date. Another interesting source of legitimacy of SBOMS was the recognition it receives from the Syrian doctors in the diaspora **(Figure 3)**.

**Figure 3:**
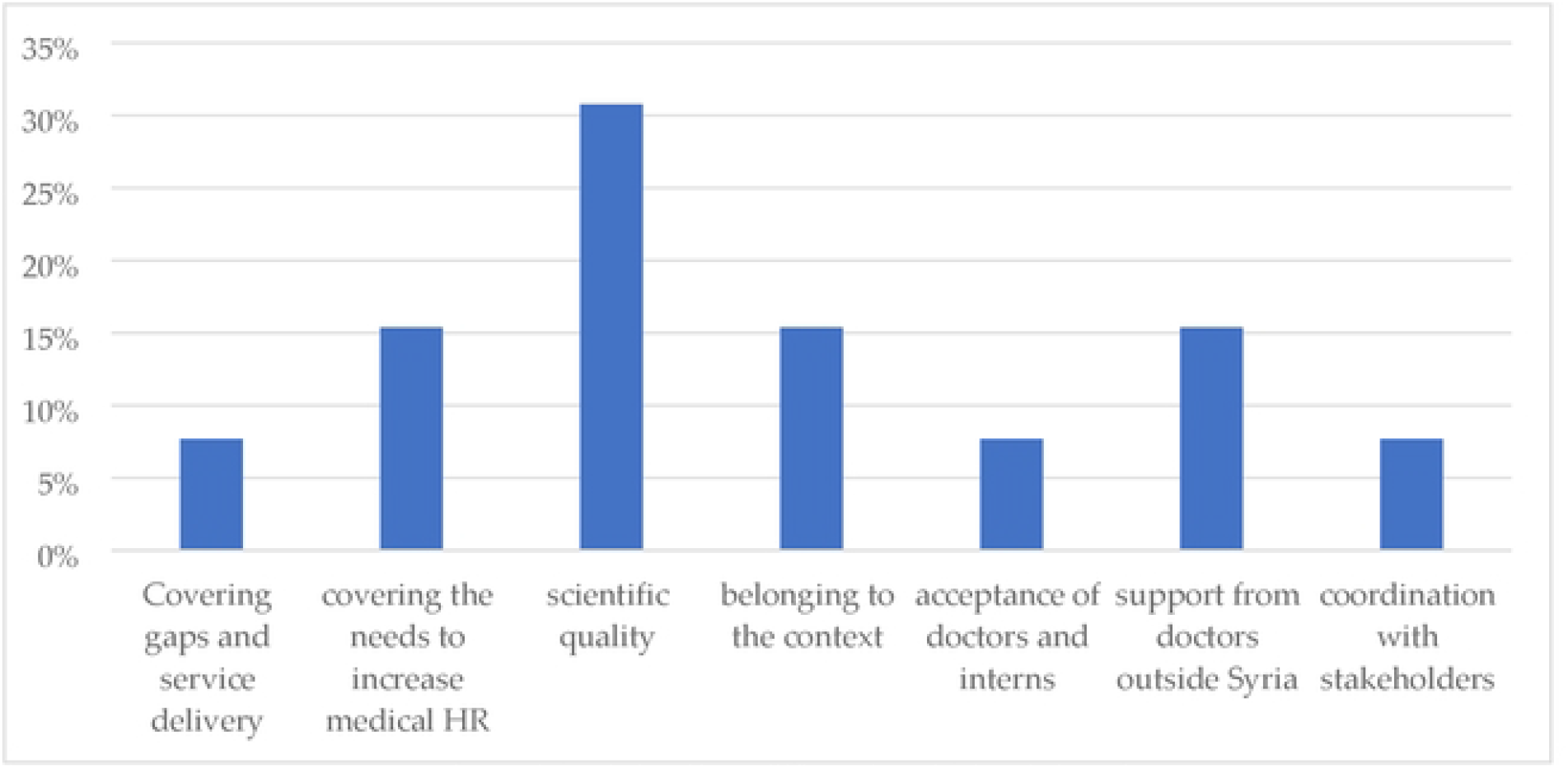
SBOMS current sources of legitimacy

SBOMS main roles include: (1) producing skilled specialized doctors; (2) developing rigorous and up to date training programs and materials for interns and medical students; (3) developing systems for examination and certification for medical graduates; and (4) monitoring and evaluating the medical training facilities in NWS. Based on these functions, the outcomes expected from the SBOMS include strengthening the capacity of health personnel in NWS, and reaching local and international recognition for SBOMS certification.

Dr Aref Razouk, stated *“The goal of the SBOMS Unit is to supply the health labour market with more specialized doctors through rigorous medical training programs and standardised examination processes. In addition, the SBOMS Unit should work to strengthen capacities in NWS for all health personnel and set standards for medical training facilities”*.

The operational model of SBOMS is focused on its educational aspects with critical coordination role among all training health facilities. Most executive activities within the training health facilities are left to the NGOs or the health directorate that run these facilities. This includes all human resources management for resident doctors and the day to day supervision.

Similar to other central bodies in NWS, the accountability system of SBOMS is still immature. This is due to the limited capacity and resources, conflict of interest among some stakeholders that play both implementation and monitoring roles within SBOMS, and the lack of interest among NGOs and potential stakeholders to develop SBOMS programs further. The decision-making mechanism in SBOMS is not completely systemized and must be developed along with the entire governance of the unit.

### Case Study 3: Infection Protection and Control (IPC) Program

Healthcare workers face numerous challenges in conflict settings, some of which are related to insufficient personal protective equipment (PPE), and ineffective infection prevention and control (IPC) practices [31]. In NWS, the idea of having a central desk for IPC emerged in 2016 by IHD, several Syrian NGOs and WHO. One of these NGOs took the lead in setting up a platform for this initiative. Accordingly, IPC guidelines and protocols were developed, many staff were trained on these protocols, and many facilities were equipped with the standard IPC infrastructure and supplies.

During the COVID-19 pandemic, the IPC program contributed to developing the outbreak response strategy. Triage tents were set up in front of most health facilities to ensure that suspected patients do not cause further transmission and that routine health services are not disrupted. The pandemic also motivated health actors to update the existing IPC guidelines and protocols to be more up to date and evidence based. Furthermore, over 500 medical personnel were trained on IPC during the COVID-19 pandemic in NWS [32].

According to the respondents’ views, the main sources of legitimacy of the IPC program are; the health directorates, and the quality of the unit’s services. SIG MoH, NGOs, and field health facilities were among the other mentioned sources of legitimacy **(*Figure 4*)**.

**Figure 4:**
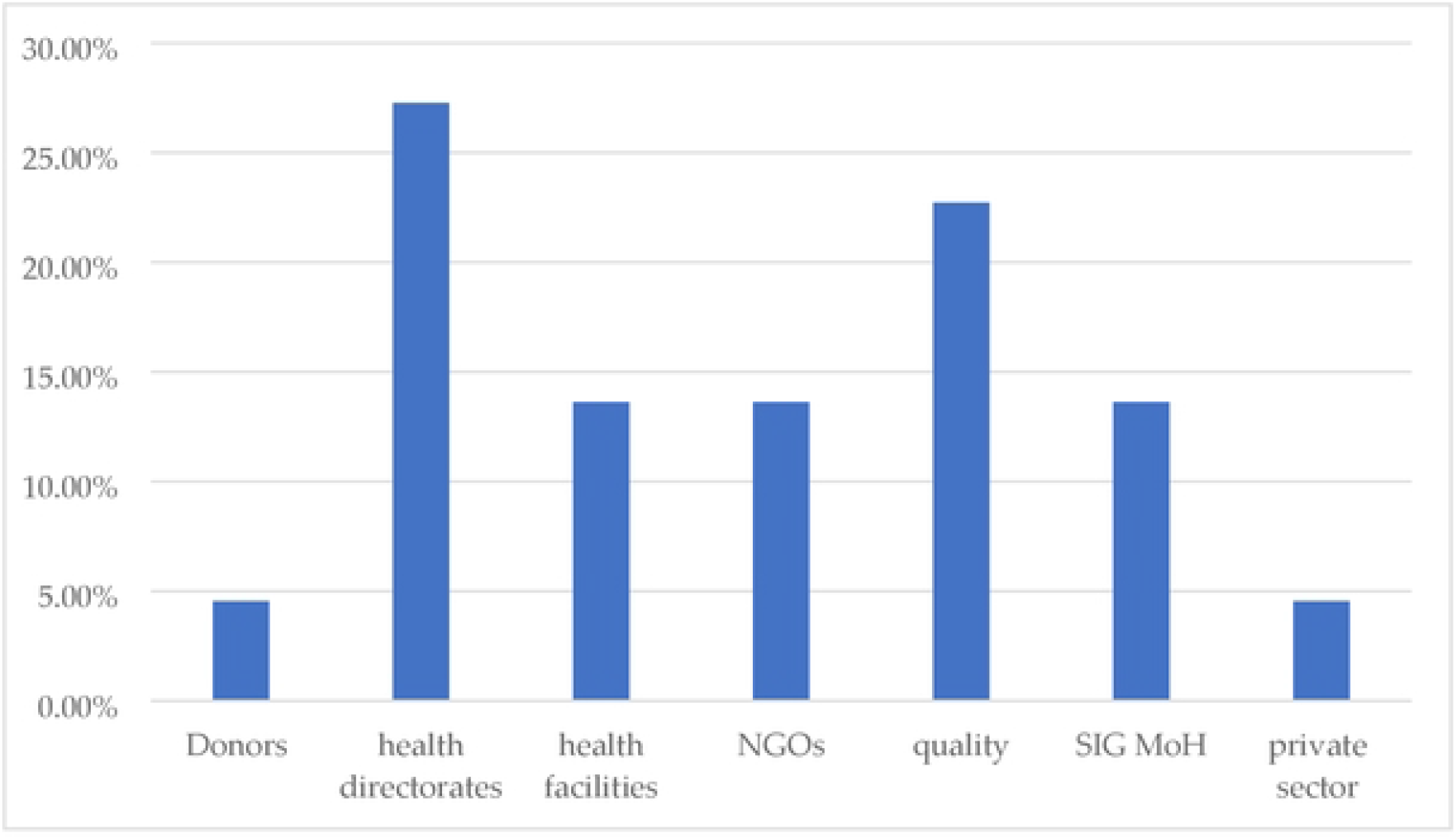
IPC unit source of legitimacy

The current roles of the IPC program, according to the participants’ views are technical and coordinative. The technical roles are related to developing IPC standardized protocols and policies, training health staff on these protocols, and improving the quality of the IPC practices. The coordination role of the unit is related to its ability to ensure collaboration from as many health actors as possible including health authorities and private sector, which would increase the geographical coverage of the program. The expected outcomes of the IPC program, accordingly, is to contribute to reducing nosocomial infections, protecting health personnel, and reducing incidence of Anti-Microbial Resistance.

The operational model of the IPC program is slightly different from the other central desks in NWS as the program is not fully independent. The IPC program is operated by a Syrian NGO semi autonomously in corporation with WHO. While the implementation of the IPC program is limited to technical aspects, the operational details related to IPC practices are left to the targeted NGOs by the IPC programs through their field health facilities. Similarly, the accountability mechanism of the program is part of the operating NGO system, which is mainly derived from donor’s compliance requirements. While the findings of the monitoring field visit -to the health facilities- are shared with a few partners separately, the practiced accountability by the NGOs is weak and needs to be developed. Moreover, the IPC program is not accountable to the health directorates or SIG MoH, which is considered by the participants as a gap in the accountability.

One of the participants stated *“The current accountability that the IPC program is that of the donor compliance requirement. The existing governance structure of the IPC Program is project-based within the overall structure of the hosting NGO. The head of programs manages the IPC Unit project manager, who manages field managers.”*

### Case study 4: The central desk for referrals

The presence of functional referral system is critical during any emergency response. Such referral system should ensure reliable links between all levels of healthcare and between the community and health facilities [33] [34]. While a comprehensive central referral system in NWS does not exist [35] [36], there were several initiatives to establishing a central referral system in NWS. According to the head of the current referral unit in NWS during the FGD sessions. “*Initially before 2016, there was a technical working group for referral and ambulance with a noticeable presence and interaction. However, it was then merged into the primary and secondary health care technical working group. As a result, the coordination was greatly affected. To overcome this shortfall, the referral central desk in NWS was established in 2016 as an initiative by a group of NGOs. In the beginning, the referral central desk it was limited in its geographical coverage due to the limited resources. Later, more stakeholders were involved, and the central desk took more coordinative role expanding its geographical coverage through collaborative partners”. During the past two years of operations (2020-2021) the central ambulance system provided medical transportation services for 87 thousand patients while the central referral team coordinated 110 thousand referrals. The most significant achievement was unifying the referral forms and registers in NWS and applying DHIS2 as the official platform for organizing and completing referrals in both Idilb and Aleppo governorates*.

The referral unit retains a high legitimacy according to 67% of the participants because of its rigorous links with the anticipated sources of legitimacy in the context of NWS **(Figure 5)**.

**Figure 5:**
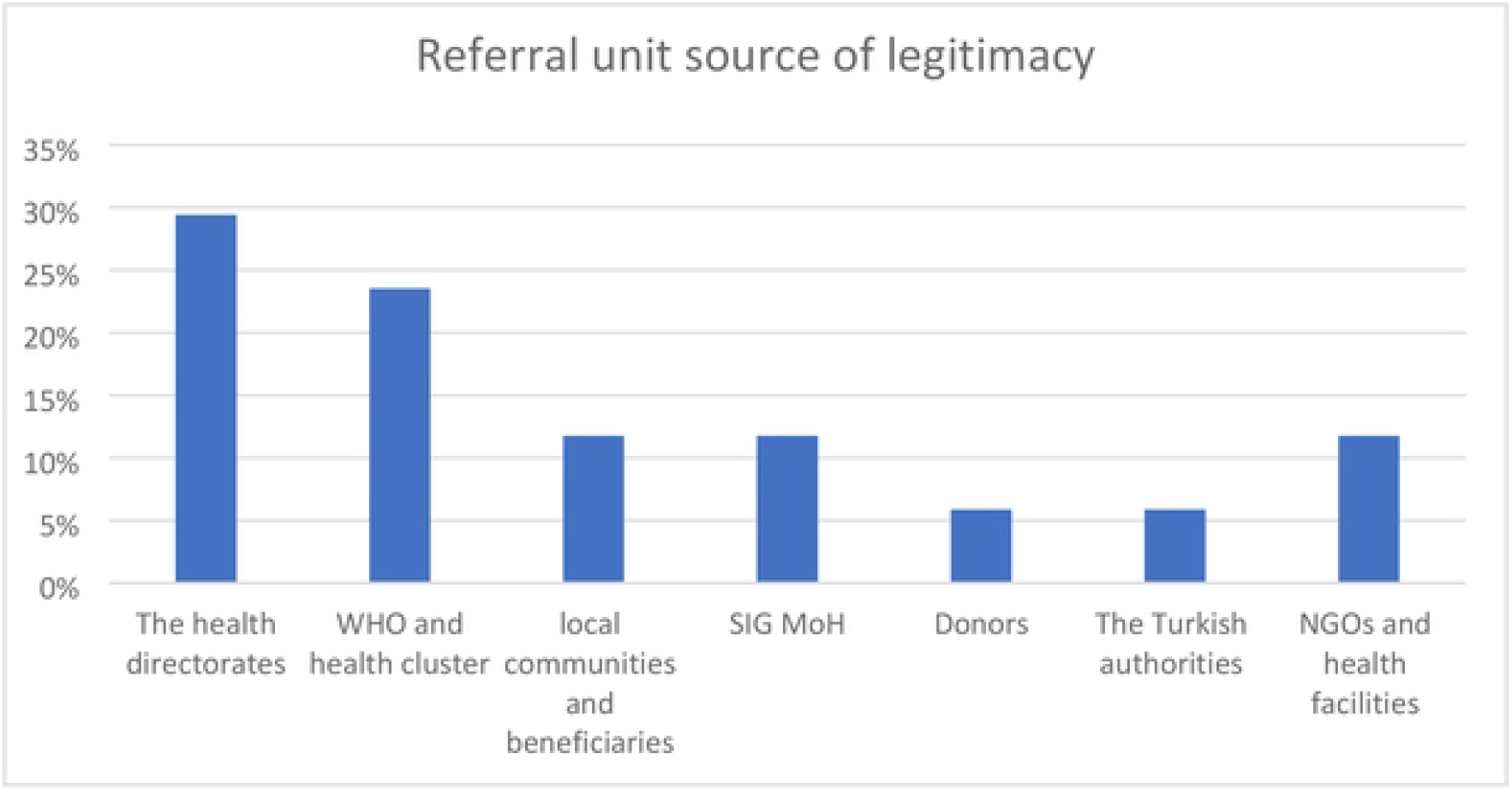
Referral unit sources of legitimacy

The main functions of the referral central desk include: (1) conduct needs assessment of ambulance and referral mechanisms, (2) develop a database of and establish connections between participating health facilities, (3) lead on communication for referral cases, (4) developing technical guidelines and procedures, and (5) build local capacities in relation to medical referrals. Additionally, the participants confirmed that the referral unit must focus more on the coordination role to engage with more health actors, which can increase the efficiency of the referral mechanisms.

The operational model of the referral unit is mainly coordinative as the unit does not have any ambulances directly owned by the unit. The unit has a leadership team in Gaziantep who work closely with the health cluster and with the NGOs, and a field team that is seconded to Idlib Health Directorate to manage its large ambulance network.

The accountability system of the referral unit is questionable based on the respondents’ answers. 25% of the participants said that the accountability mechanism of the unit is poor. According to the responses, there is a lack of clarity about the position of the central referral Unit, its leadership, and its reference to the NGOs and the HDs that host this unit. Other participants argued that this ambiguity is related to the challenges facing the unit and the uncertainty in its structure and hierarchy. Some of these challenges are related to lack of clarity in the governance of the health system with many gaps and complications in the presumed governance system. For the referral central desk, this lack of clarity affect its ability to position it self within the health sector.

For accountability, there is project-based partial accountability to WHO, the health cluster, and the technical working groups. The referral desk produces periodic reports that are shared with these entities. However, there is a lack of information about service quality and beneficiaries’ satisfaction. Therefore, there is a defect in the transparency of the referral Unit.

## Discussion and conclusion

Considering the complicated health governance in northwest of Syria, health actors had to come up with new approaches to navigate through conflict sensitises and complex geopolitics. Central desks approach is one of the innovative approaches that have been used in northwest Syria and proved some levels of efficiency. Our paper found that the use of central desks in northwest Syria is believed to have supported the health sector in the region and contributed to better health outcomes.

These findings are in line with the limited literature on central desks/projects approach. During the Ebola outbreak between 2014 and 2018 there was a need to centralise health information and patient records to ensure complementarity, avoid duplications, and avoid interruption of treatment. Some actors implemented central Health Information System (HIS) in the region that improved the availability of health information, and consequently allowed health actors to have better planning and strategies. A study that assessed the functionality and the effectiveness of the central HIS that was developed during the Ebola response in Sierra Leone and found significant improvements in the availability of health records. They found that after this central project, all patients had basic records (100%, n = 456/456), and the majority of patients had records with details on medications, clinical presentations and epidemiological records (98.9%, n = 451/456) [11]. Another study looked at some tech interventions to strengthen electronic records systems on district level in Sierra Leone. The study concludes that implementing mobile health interventions to support the surveillance system of Ebola improved data completeness, storage and accuracy [12]. These examples support the argument that technical central structures with locally developed solutions can improve health related outcomes.

In the case of northwest Syria although central desks vary widely in their role, structure, vision, and operational modality, they do share some common features. Our paper found that the key features that made the central desks an effective governance approach can be summarised as follow. First, independence and autonomous structure, which allowed the various health actors to feel comfortable engaging and coordinating with these desks without the fear of political implications. Second, technical focus on a specific issue in the health sector, which allowed these central desks to establish their specific niche and avoid being perceived by other health actors as competitors. Third, flexibility to work with the various types and levels of actors including local health authorities, interim government, humanitarian NGOs, UN agencies, and diaspora networks.

With regards to legitimacy of central desks in northwest Syria, our paper found that these desks derive their legitimacy mainly from the critical gap they fill in the health sector, the quality of their outcomes and products, and their ability to work closely with the health directorates from the one hand and with the NGOs and other humanitarian actors from the other hand.

Central projects represent an innovative approach for health governance in chronic conflict settings. While health governance generally is a neglected issue in emergency responses, humanitarian actors should pay more attention to health governance approaches and tools especially in chronic crises. The traditional approaches of many humanitarian actors have an underlying assumption that crises short-termed, which is not the case in almost all recent conflicts. As a result, humanitarian health actors are usually highly skilled in relation to life saving and short-term emergency responses, but they usually fall short in sustainability and early recovery where they should focus on issues like HIS, health workforce development, IPC, to pave the way for development interventions. On the other hand, traditional development interventions have an underlying assumption that there is a legitimate national government that can lead development projects. However, it is not uncommon to have areas that are not governed by a legitimate government. Countries suffering from a civil war, such as Syria, Libya, Yemen, and Somalia, have large territories controlled by militia; many of them might be prescribed as terrorist groups, further complicating service provision. Humanitarian and development actors cannot wait until a legitimate government emerge in such settings before they start joining efforts in tackling shared challenges and strengthening local systems.

## Data Availability

Data cannot be shared publicly because of conflict sensitivity. The data is related to the health governance in northwest Syria, which is a contested conflict settings. Nevertheless, the anonymised analysis of the Focus Group Discussions can be found on: DOI:10.17632/9k69zkmg3z.1

https://doi.org/10.17632/9k69zkmg3z.1

## Acknowledgement

- *Dr. Rami KALAZI, Dr. Okba DOGHIM, Dr. Mossab BREIJ; for their great contribution to the FGD note transcribes*.
- *Dr. Ahmed Kalecioglu, Dr. Amer Heluani, Dr. Mahmoud HARIRI, Dr. Safwan Alchalati*, Dr Salah Safadi; for their substantial contribution for the validation of key findings of the FGD.

